# Incidence of Chronic Recurrent Multifocal Osteomyelitis in Children and Adolescents in the UK and Republic of Ireland

**DOI:** 10.1101/2024.06.06.24308477

**Authors:** Daphne Theresa Chia, Andoni Toms, Anish Sanghrajka, Athimalaipet Ramanan, Orla G Killeen, Cristina Ilea, Kamran Mahmood, Sandrine Compeyrot-Lacassagne, Kathryn Bailey, Neil Martin, Kate Armon, Chenqu Suo

## Abstract

**Objectives:** Chronic Recurrent Multifocal Osteomyelitis (CRMO), also known as chronic nonbacterial osteomyelitis (CNO), is a rare autoinflammatory condition affecting the bones in children and teenagers. The actual incidence of CRMO remains uncertain. The objective of this study is to identify the incidence of CRMO in children and young people under the age of 16 years in the United Kingdom (UK) and Republic of Ireland (ROI). We also aim to delineate the demographics, clinical presentation, investigations, initial management and healthcare needs for children and adolescents with CRMO.

**Methods:** We conducted monthly surveys among all paediatric consultants and paediatric orthopaedic surgeons to identify patients newly diagnosed with CRMO between October 2020 and November 2022. A standardised questionnaire was sent to reporting clinicians to collect further information.

**Results:** Over the surveillance period, 288 patients were reported, among which, 165 confirmed and 20 probable cases were included in the analysis. The highest incidences were among 8-10 year-olds. A two-to-one female-to-male difference in incidence was observed, and male patients were more likely to present with multifocal disease. A negative correlation was observed between reporting clavicular and leg pain. Investigation-wise, 80.0% of patients were reported to have undergone whole-body MRI and 51.1% had bone biopsies. The most common initial treatments were NSAIDs (93.9%) and bisphosphonates (44.8%).

**Conclusion:** This study estimates an average annual CRMO incidence of 0.65 cases per 100,000 children and adolescents in the UK and ROI. These findings establish a crucial baseline for ongoing research and improvement in the care of individuals with CRMO.

**Key Messages:**

- The estimated annual incidence of CRMO is 0.65 per 100,000 children and adolescents aged <16.
- Two-fold higher incidence in females to males was found, with the highest incidence in 8-10 year-olds.
- Male patients are more likely to present with multifocal disease compared with females.

## Introduction

Chronic Recurrent Multifocal Osteomyelitis (CRMO), also known as chronic nonbacterial osteomyelitis (CNO), is a rare autoinflammatory condition affecting the bones. It occurs primarily in children and teenagers and is characterised by bone pain and swelling in the absence of infection or tumour.

Though first described over five decades ago [1], the true incidence of CRMO in children and adolescents remains uncertain. Previous studies [2–6] have focused on patients identified through rheumatology centres. The only nationwide surveillance study to date is the German Surveillance Study (2006-2008) which estimates an incidence of 0.4/100,000/year [7]. Notably, no national study has ever been carried out in the United Kingdom (UK) and Republic of Ireland (ROI) population to look at the disease. Studies report a delay in diagnosis due to under-recognition [2], and there are no randomised controlled trials for treatment.

To address these knowledge gaps, we initiated a prospective surveillance study through the British Paediatric Surveillance Unit (BPSU) and British Society for Children’s Orthopaedic Surgery (BSCOS). Our primary objective is to establish the incidence of CRMO in children and adolescents aged under 16 in the UK and ROI. Additionally, we aim to comprehensively investigate the demographics, clinical presentation, time to diagnosis, investigations, initial management and healthcare needs of patients with CRMO.

## Methods

### Study design

A prospective surveillance study was undertaken from October 2020 to November 2022. The BPSU sends a monthly orange eCard to all registered paediatric consultants in the UK and ROI. Since 1985, BPSU has successfully facilitated research in more than 100 rare conditions in children [8]. Through the BPSU orange eCard, we asked if registered paediatric consultants have seen any patients (up to but not including age of 16 years) with a new diagnosis of possible CRMO, according to our surveillance case definition. A detailed questionnaire was sent to the reporting clinicians to collect further information about each case. Clinicians returned patient identifying details via a password protected PDF form, and clinical details via either a secure electronic system (REDCap) or password protected PDF. A parallel surveillance study was run through the BSCOS to identify patients who were managed by only paediatric orthopaedic surgeons without any input of a paediatrician.

### Case definition

Clinicians were asked to report new cases in the past month which met the surveillance case definition:

Children and young people up to but not including the age of 16 years with a new diagnosis of possible CRMO, namely those who have the following features:

- The presence of localised bone pain, which could be single site or multiple sites

AND

- The presence of typical radiological findings on plain X-ray (examples include: lysis, sclerosis, cortical thickening or periosteal reaction) or on MRI (examples include: bone marrow oedema on fluid sensitive sequences, or periostitis (periosteal inflammation))

AND

- The treating clinician has determined that the clinical features are not explained by an alternative diagnosis e.g. trauma, infection or neoplasm

The surveillance case definition was written to be more inclusive than the analytic case definition, to ensure all potential cases are reported. The analytic case definition (Supplementary 1A) was based on the Bristol Diagnostic Criteria [2]. Cases that might suggest alternative diagnoses were excluded: children with low ALP (might indicate hypophosphatasia) or children with positive blood culture or positive culture on bone biopsy (might indicate infective cause). Complex cases were discussed in the study management group to determine by consensus whether they met the analytic case definition. The Bristol criteria were developed primarily to determine when children should undergo bone biopsy for confirmation of the diagnosis / exclusion of other conditions and were published at a time when whole body MRI was less available. Therefore, we have included a category of ‘probable CRMO’ for cases that did not meet the analytic case definition, but the study management group agreed that it was highly likely to be CRMO, and appropriate investigations were carried out to exclude other conditions. A decision tree illustrating the case inclusion and exclusion criteria is shown in Supplementary 1B.

### Data analysis

Cases were de-duplicated using unique identifiers such as NHS and CHI numbers. Confirmed CRMO cases and probable CRMO cases were included in our analysis. Incidence was expressed as the number of cases per 100,000 person-years. The data for the total number of children <16 in 2020-2022 was obtained from:

● Average of the Office of National Statistics mid-2020 and mid-2021 population estimates for children aged <16 in the UK.
● Central Statistics Office 2022 population census for children aged <16 in the ROI.

### Statistical analysis

Statistical analysis was performed using R (Version 4.2.1). Numeric data was expressed as median and interquartile range (IQR: 25th, 75th percentile). When comparing the male and female groups, Wilcoxon rank sum test was used for continuous data and Pearson’s chi-square test for categorical data. If the expected square count was less than five, a Fisher’s exact test was used. When comparing the incidence across different ethnicities, data on ethnicity distribution among children aged 5 to 16 in the UK in 2020 [9] was used, hence cases from ROI and those <5 years were excluded from the CRMO dataset. Fisher’s exact test was used to calculate the p-value. In the multivariate logistic regression analysis, cases with missing data in any of the tested covariates were excluded. P-values were adjusted for multiple testing using the Benjamini-Hochberg procedure. P-values less than 0.05 were considered statistically significant.

### Ethical approval

This study was approved by the BPSU Scientific Committee. Ethical approval was gained from London - Central Research Ethics Committee (reference: 20/LO/0195); HRA Confidentiality Advisory Group (reference: 20/CAG/0029); Public Benefit and Privacy Panel for Health and Social Care (reference: 1920-0202).

## Results

### Reported cases and estimated incidence

From October 2020 to November 2022, 282 newly diagnosed CRMO cases were reported from the BPSU orange e-reporting card (Figure 1). Of the cases reported from BSCOS, 6 had no paediatric involvement after deduplication. Out of the 288 initially reported cases, 91% had case questionnaires returned.

**Figure 1:**
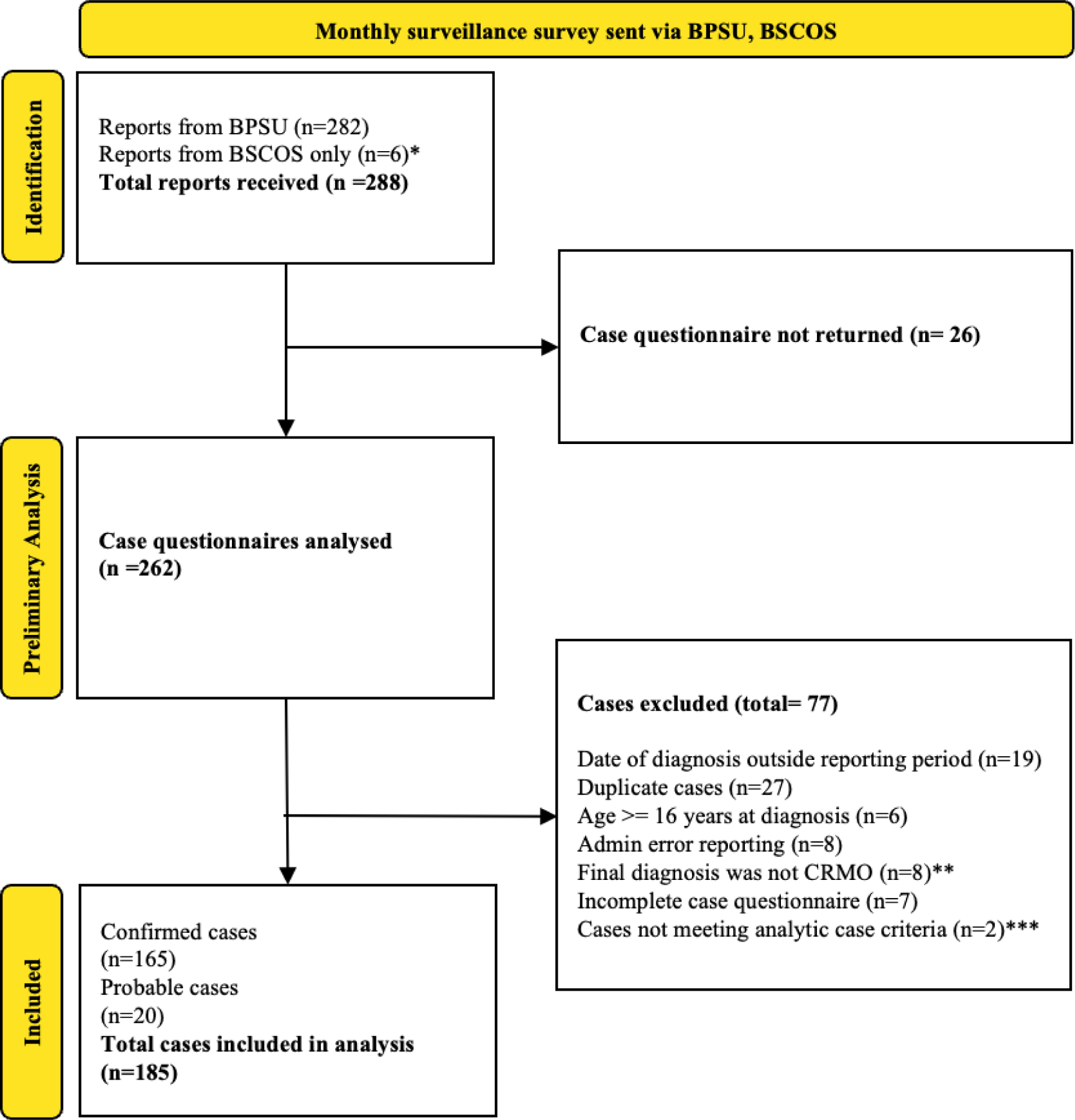
Schematic summary of reported cases from BPSU & BSCOS. *BSCOS reported cases which had no paediatric involvement (n=6) **These cases were initially reported as suspected CRMO but diagnoses were changed on subsequent correspondence with the reporting physicians. The final diagnoses were: 1 Langerhans cell histiocytosis, 1 stress fracture, 1 post traumatic thoracic vertebral changes, 1, idiopathic juvenile osteoporosis, 4 unknown. ***Both cases were discussed in the study management group, one was in keeping with infectious osteomyelitis (CRP 81, MRSA in multiple blood cultures and images in keeping with infectious osteomyelitis), one excluded as no evidence of any abnormality on imaging.

In total, 165 children and young people with confirmed CRMO and 20 with probable CRMO were reported over the 25-month surveillance period and were included in the analysis. The calculated incidence of CRMO for children under 16 in the UK and ROI was 0.65 per 100,000 person-years. Using the abbreviated postal code of each reported case, we calculated the estimated incidences in the 12 areas in the UK together with ROI (Supplementary Figure S2) and found that the estimated incidences range from 0.12 to 1.32 cases per 100,000 person-years.

### Demographics and Clinical Features

The median age at the onset of symptoms was 9.5 years (IQR 8.0, 11.0) (Table 1, Figure 2A), while the median age at the time of diagnosis was 10.2 years (IQR 8.8, 12.2). The calculated incidences by age are shown in Supplementary Figure S3, with the highest incidences in 8 to 10 years old. The median duration from symptom onset to diagnosis was 5 months (IQR 3, 12). Notably, 26.6% (46/173) of patients experienced a delay in diagnosis greater or equal to 12 months (Supplementary Figure S4). Two-thirds (65.4%) of patients were female. The calculated incidence in males (<16 years) was 0.44 per 100,000 person-years. The calculated incidence in females (<16 years) was 0.88 per 100,000 person-years. Ethnicity was recorded according to the Office for National Statistics 2011 Coding for Ethnic Groups. A significant majority of patients were identified as White (158/183, 86%), while 11 (6.0%) were of Asian or British Asian ethnicity, 9 (4.9%) were of mixed or multiple ethnic background, and 4 (2.2%) were of Black, African, Carribean or Black British Background. The incidence of CRMO was significantly higher in White ethnicity compared to other ethnicities in the general UK population (p-value 0.031).

**Figure 2:**
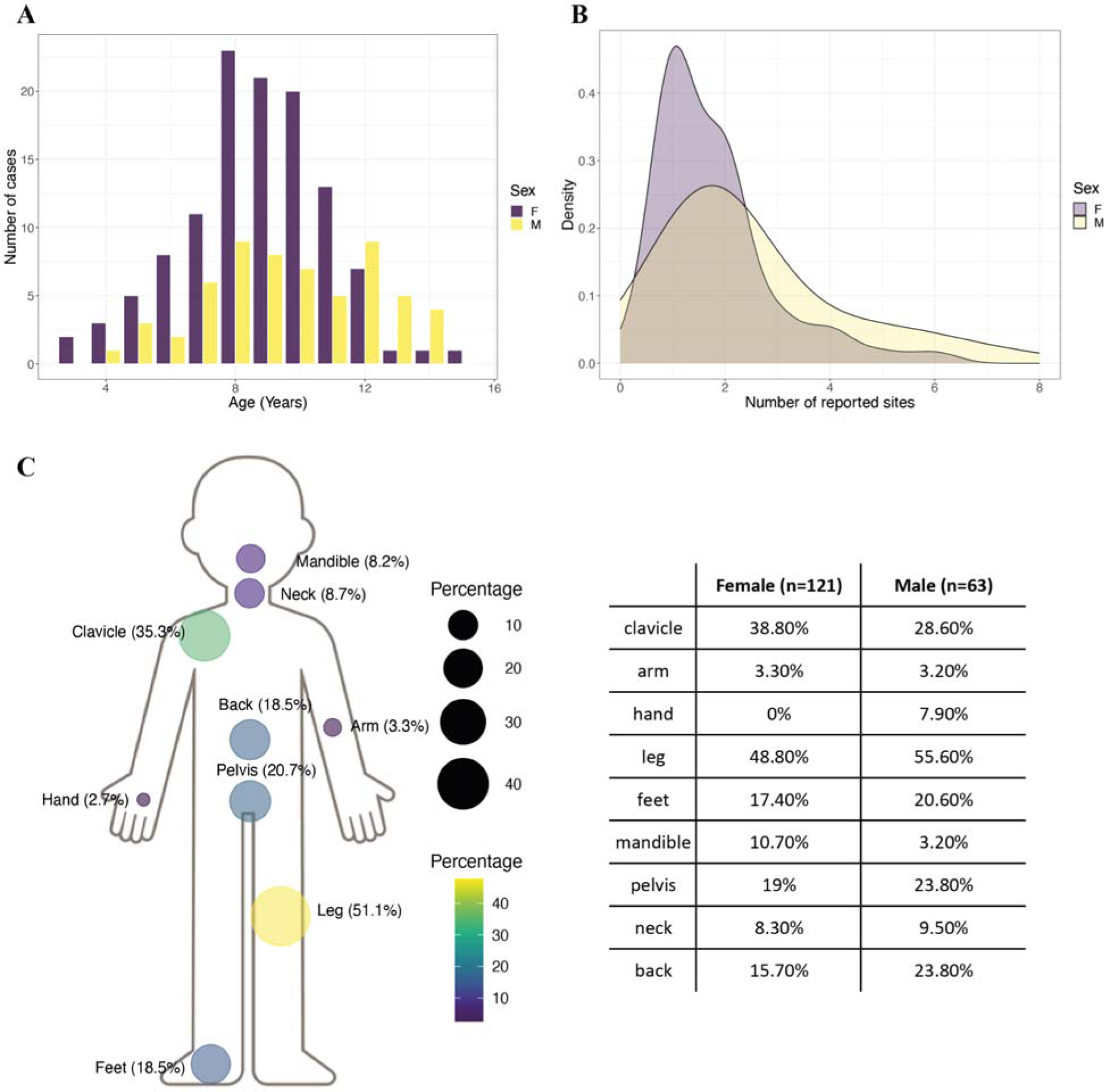
Age of onset and number of reported painful sites. (A) Density graph of age at onset of symptoms, comparing male and female patients. (B) Density graph of number of reported painful sites per patient, comparing male and female patients. (C) Body map of reported sites of bone pain (left), with table of reported sites of bone pain split by gender (right).

**Table 1:** Demographic and clinical features at presentation.

| Characteristic | Overall<br>n = 185 | Female<br>n = 121 | Male<br>n = 64 | p-value (adjusted) |
| --- | --- | --- | --- | --- |
| **Age at onset** |  |  |  | 0.038<sup>1</sup> (0.139) |
| **Median (IQR)** | 9.5 (8.0 - 11.0) | 9.3 (8.0 - 10.7) | 10.1 (8.1 - 12.3) |  |
| **Range** | 3.5 - 15.1 | 3.5 - 15.1 | 4.2 - 14.7 |  |
| **Time to diagnosis (months)** |  |  |  | 0.6<sup>1</sup> (0.73) |
| **Median (IQR)** | 5 (3 - 12) | 6 (3 - 13) | 5 (3 - 10) |  |
| **Clinician reported sites** |  |  |  | 0.007<sup>2</sup> (0.044) |
| **Multifocal** | 94 (57%) | 55 (50%) | 39 (72%) |  |
| **Unifocal** | 70 (43%) | 56 (50%) | 15 (28%) |  |
| **Ethnicity** |  |  |  | 0.3<sup>3</sup> (0.55) |
| **White** | 158 (86%) | 104 (87%) | 54 (86%) |  |
| **Asian/Asian British** | 11 (6.0%) | 9 (7.5%) | 2 (3.2%) |  |
| **Mixed/Multiple** | 9 (4.9%) | 4 (3.3%) | 5 (7.9%) |  |
| **Black/African/**<br>**Caribbean/Black British** | 4 (2.2%) | 3 (2.5%) | 1 (1.6%) |  |
| **Other Ethnic Group** | 1 (0.5%) | 0 (0%) | 1 (1.6%) |  |
| **Bone Swelling** | 111 (60%) | 81 (67%) | 30 (47%) | 0.008<sup>2</sup> (0.044) |
| **Bone Deformity** | 15 (8.1%) | 9 (7.4%) | 6 (9.4%) | 0.6<sup>2</sup> (0.73) |
| **Joint Swelling** | 38 (21%) | 21 (17%) | 17 (27%) | 0.14<sup>2</sup> (0.308) |
| **Skin Lesions** | 26 (14%) | 16 (13%) | 10 (16%) | 0.7<sup>2</sup> (0.77) |
| **General Malaise** | 27 (15%) | 16 (13%) | 11 (17%) | 0.5<sup>2</sup> (0.80) |
| **Fever** | 19 (10%) | 13 (11%) | 6 (9.4%) | 0.8<sup>2</sup> (0.80) |
| **Weight Loss** | 13 (7.0%) | 6 (5.0%) | 7 (11%) | 0.14<sup>3</sup> (0.31) |

<sup>1</sup> Wilcoxon rank sum test
<sup>2</sup> Pearson's Chi-squared test
<sup>3</sup> Fisher's exact test

Clinicians were asked to report the presenting clinical features of each patient (Table 1). The median number of reported painful sites was 2, with 43% (70/164) presenting with unifocal bone pain.

Male patients were significantly more likely to present with multifocal disease (39/54, 72%) compared with female patients (55/110, 50%) (Table 1, Figure 2B). Conversely, significantly more females presented with bone swelling (81/121, 67%) compared with males (30/64, 47%).

Figure 2C illustrates the distribution of reported bone pain at presentation. The most common site of bone pain was in the leg (94/184, 51.1%), followed by clavicle (65/184, 35.3%), pelvis (38/184, 20.7%), back (34/184, 18.5%) and feet (34/184, 18.5%). In addition, 15 patients presented with pain in the mandible (8.2%). There was no significant difference between reported sites of pain between females and males (Figure 2C). Unilateral clavicular bone pain was more common (59/65, 90.8%) than bilateral. The reporting of bone pain in the clavicle demonstrates a significant negative correlation with the reporting of bone pain in the leg (Pearson correlation coefficient -0.32, adjusted p-value 0.00028). Additionally, there was a significant negative correlation (though less marked) between leg and mandible (Pearson correlation coefficient -0.23, adjusted p-value 0.038). Patients with reported pain in the clavicle but not leg are more likely to present with unifocal disease compared to multifocal disease pattern for those with reported pain in the leg but not clavicle (Supplementary Figure S5A). This suggests two patterns of symptomatic presentation, one characterised by unifocal disease in the clavicle and another exhibiting a multifocal pattern affecting the legs along with other sites.

Associated personal medical history included psoriasis (12/162, 7.4%), inflammatory bowel disease (6/162, 3.7%) and palmoplantar dermatitis (4/162, 2.5%). In first-degree family members, psoriasis was the most commonly reported (23/162, 14.2%). This is five times the estimated prevalence of psoriasis in the UK [10]. This was followed by rheumatoid arthritis (7/162, 4.3%), inflammatory bowel disease (6/162, 3.7%), thyroid disease (6/162, 3.7%), coeliac disease (3/162, 1.9%) and other autoimmune diseases (5/162, 3.1%). A family history of CRMO was suspected in 3 patients and confirmed in only 1 patient.

### Investigations

A median of 3 imaging tests were performed per patient. Figure 3A shows the proportion of patients investigated with each modality. C-reactive protein (CRP) was reported for 178/185 (96.2%) of patients. Figure 3B shows the distribution of CRP values, with 86.5% (154/178) having CRP below 30.

**Figure 3:**
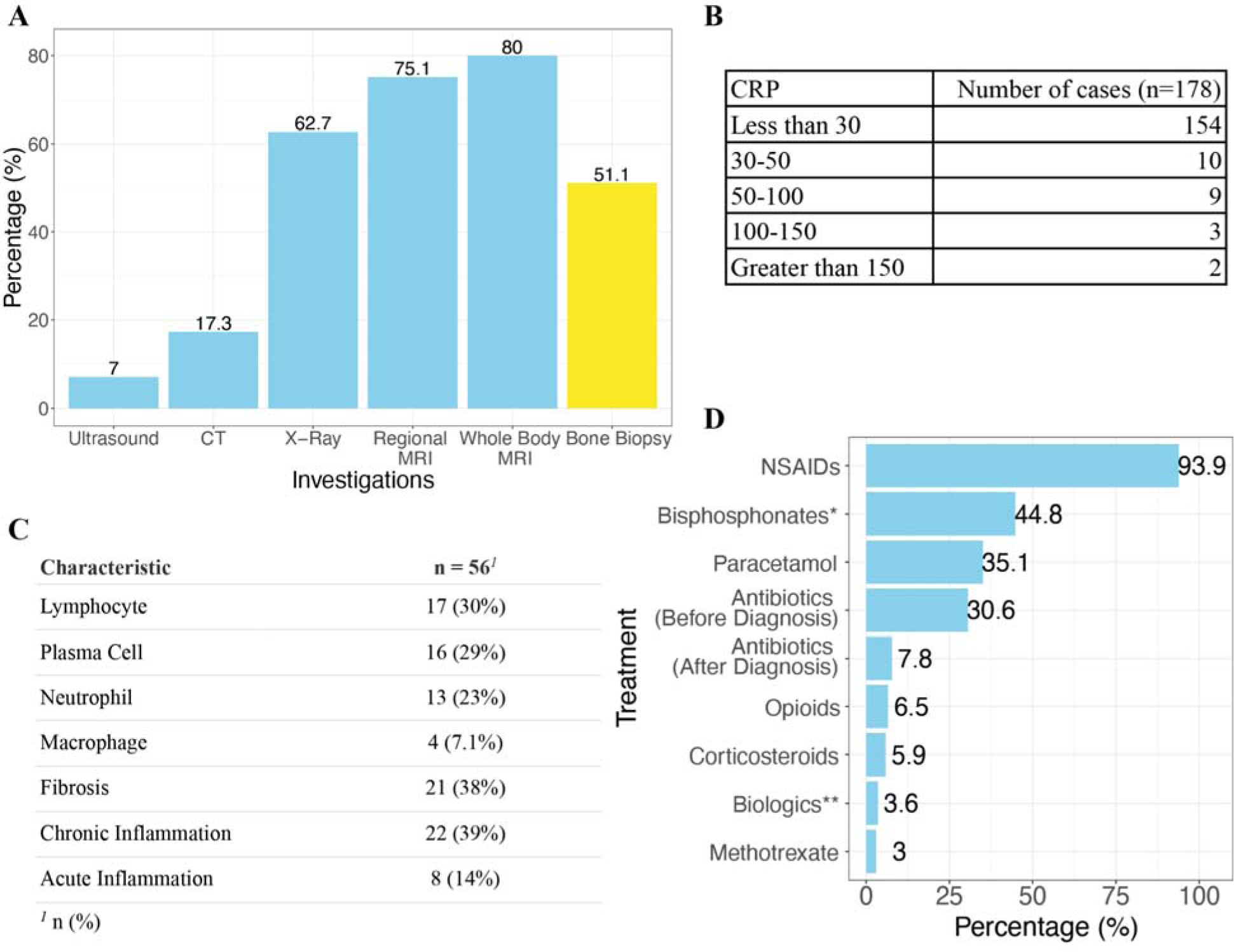
Investigations and Management. (A) Bar chart showing the percentages of patients having had different imaging modalities and bone biopsy. (B) Table of reported CRP values. If the test had been done more than once at the time of reporting, the clinician was asked to record the highest value. (C) Analysis of positive histopathology characteristics from the bone biopsy reports received (n = 56). (D) Treatment received at the time of reporting *Bisphosphonates: Of 72 patients reported to use bisphosphonates, pamidronate = 58, zoledronate = 14 **Biologics: Of 6 patients reported to use biologics, vedolizumab = 2, etanercept = 1, infliximab = 1, ustekinumab = 1, adalimumab = 1

Sixty-nine patients (69/135, 51.1%) had bone biopsies, of which 56 biopsy reports were submitted. Bone biopsy reports were analysed using word search, where pathological terms with similar meanings were grouped for analysis (Figure 3C). Nine showed no features, 17 had a single feature and 30 had more than one feature. Chronic inflammation (n=22/56, 39%) and fibrosis (n=21/56, 38%) were the most common descriptions. The most common cell types described were lymphocytes (n=17/56, 30%) and plasma cells (n=16/56, 29%). The relationship between the histopathology features is shown in a chord diagram (Supplementary Figure S6).

### Initial Management

The median time interval from date of diagnosis to date of completion of the questionnaire was 3 months (IQR 2, 7). Figure 3D shows the treatment received by the patients at the time of reporting, which was therefore within a few months of diagnosis. The most common treatment was NSAID (168/179, 93.9%). This included ibuprofen (75/179, 41.9%), naproxen (66/179, 36.9%), piroxicam (6/179, 3.4%) and diclofenac (3/179, 1.7%). Bisphosphonates were also commonly used (78/174, 44.8%), including pamidronate (58/174, 33.3%) and zoledronate (14/174, 8.0%). Fewer patients received steroids (10/170, 5.9%) and biologics (6/168, 3.6%). For two on biologics, the treatment was for concurrent IBD. Methotrexate was used in 5/168 (3.0%) of patients. Prior to diagnosis, antibiotics were used in 55/180 (30.6%) of patients. After diagnosis, this proportion decreased to 14/180 (7.8%) of patients. A multivariate logistic regression analysis (Supplementary Figure S5B) revealed that younger patients (adjusted p-value 0.0093) and patients with higher CRP (adjusted p-value 0.0018) were more likely to receive antibiotics.

### Healthcare Utilisation & Patient Function

Most patients (110/184, 59.8%) were diagnosed by rheumatologists (including paediatric rheumatologists and general paediatricians with rheumatology interest) (Figure 4A). Diagnosis was made by orthopaedic surgeons in 32 patients (17.4%), and general paediatricians in 18 patients (9.8%). Seventeen patients (9.2%) were diagnosed within a multidisciplinary team (usually including paediatrics, orthopaedics, radiology and histopathology specialists). Three patients were diagnosed by maxillo-facial surgeons (1.6%) and all three presented with isolated mandibular pain.

**Figure 4:**
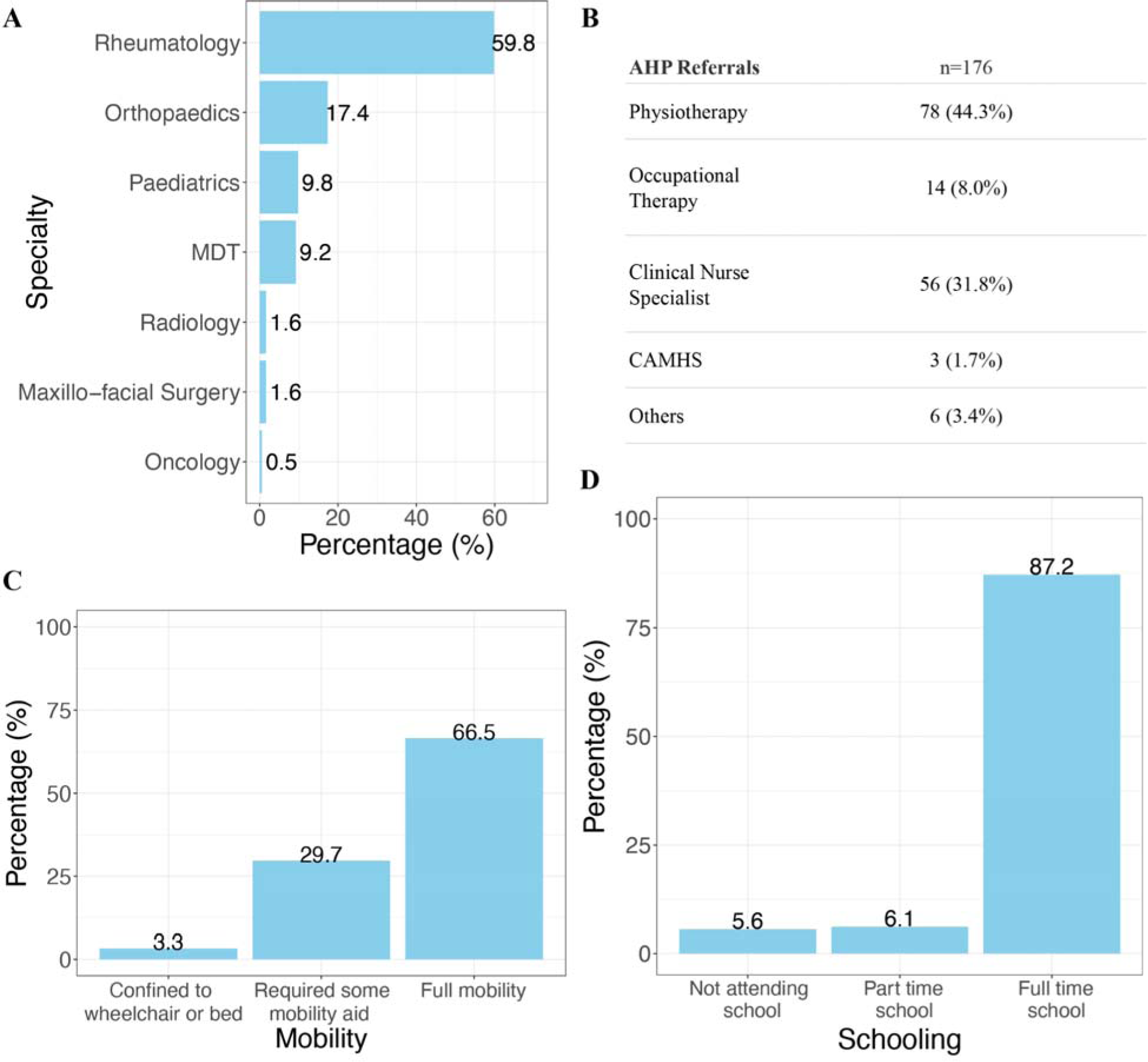
Health Utilisation & Patient Function. (A) Bar chart showing the percentage of patients with CRMO diagnosed by each specialty. (B) Utilisation of allied health professionals at time of reporting. (C) Impact of CRMO on mobility at presentation. (D) Impact of CRMO on schooling at presentation.

Figure 4B shows referrals made to allied health professionals. The most common referral was to physiotherapy (78/177, 44.1%), followed by clinical nurse specialists (56/176, 31.8%) and occupational therapy (14/176, 8.0%). Only 3 patients were referred to a Child and Adolescent Mental Health Service at the time of reporting.

When the reporting clinician first met the patient, 157/178 (88.2%) were able to attend full-time school (excluding children and young people with CRMO that were home-schooled or too young to attend school, Figure 4C), while 11/178 (6.2%) attended part-time school and 10/178 (5.6%) did not attend school. In addition, 54/182 (29.7%) required some mobility aid, 6/182 (3.3%) were confined to a wheelchair/bed, and only 121/182 (66.5%) had full mobility (Figure 4D). We note that there was a relatively high percentage of children and young people requiring mobility aid; the reporting clinicians may have included those with mild impairment in this group (though not necessarily requiring aid) as there was no separate category for this.

Prior to diagnosis, 65/174 (37.4%) of children and young people with CRMO had an inpatient admission relating to presenting features. Of these patients, the median number of admissions was 1 (IQR 1,1.5) and the median length of stay was 5 days (IQR 2,7). Accident and Emergency (A&E) attendance due to CRMO (prior to diagnosis) was recorded for 61.6% of patients (98/159). Of these patients, the median number of A&E visits were 1 (IQR 1,2). There were no deaths at time of reporting.

## Discussion

### Summary

This study is the first prospective nationwide surveillance study of CRMO in the UK and ROI. The calculated annual incidence of CRMO in the UK and ROI was 0.65 per 100,000 children (<16 years). It was more common in females (2:1 female:male) and in white ethnicity.

These findings are in line with previously reported estimates from Germany and Australia, ranging from 0.4 to 1.0 per 100,000 children per year [4,7]. Additionally, the regional incidence varies from 0.12 to 1.32 cases per 100,000 children per year. This variability may stem from factors such as under-diagnosis, under-reporting or demographic differences across regions. Furthermore, the fact that the highest incidence is reported in the East of England where the study group is based might imply that the upper limit of the true incidence of CRMO could be around 1.32 cases per 100,000 children (<16 years) per year.

This study stands as the third largest documented cohort of children and young people with CRMO (Table 2), and represents the most extensive surveillance study conducted on this disease. The unbiased nature of the surveillance enables capture of all paediatric CRMO cases under the care of either paediatric consultants or paediatric orthopaedic consultants, regardless of disease severity. This stands in contrast to potential bias towards patients with severe CRMO identified by paediatric rheumatologists or from tertiary centres. The dual surveillance of both paediatric consultants and paediatric orthopaedic consultants enables the identification of relatively uncommon cases where only orthopaedic surgeons were involved. We did not survey maxillo-facial surgeons, who could potentially diagnose and manage CRMO independently, although this number is expected to be small. In our study, maxillo-facial surgeons provided the initial diagnosis in three patients, with subsequent management by paediatric rheumatologists.

**Table 2:** Summary of CRMO studies with cohort number above 100.

| Paper | Country/region | Type of study | Source of cases | Number of patients | Time period | Age group included |
| --- | --- | --- | --- | --- | --- | --- |
| Jansson (2011) [7] | Germany | Nationwide surveillance | Paediatric and paediatric orthopaedic hospitals | 148 | 2006-2008 | 18 months - 18 years |
| Wipff (2015) [6] | France | Multicentre study | 12 centres | 178 | 1995-2014 | < 18 years |
| Bhat (2018) [5] | UK | Multicentre study | 8 centres | 131 | 2001-2017 | < 18 years |
| Girschick (2018) [3] | Mainly European countries | Registry | Eurofever registry | 286 | 2009-2016 | unlimited (including adult patients) |
| Aden (2022) [11] | USA | Single Centre study | 1 centre | 215 | 2005-2019 | < 18 years |
| This study | UK, ROI | Nationwide surveillance | paediatric and paediatric orthopaedic consultants | 185 | 2020-2022 | < 16 years |

### Clinical Presentation

The demographic features of our patient cohort aligns closely with previous studies regarding age and gender distribution [2–7,11]. The median time to diagnosis in our cohort (5 months) was shorter compared with previous studies [3–7,12,13], which may be attributed to an increased awareness of CRMO among clinicians, and increasing availability of whole-body MRI.

We found that male patients were significantly more likely to present with multifocal disease. This is consistent with the French study [6] where the severe phenotype primarily comprised male patients, most of whom exhibited a multifocal form of CRMO. In contrast, female patients in our cohort were significantly more likely to present with bone swelling, a trend that warrants further confirmation through future studies. Additionally, we identified a significant negative correlation between clavicular pain and leg pain, suggesting two possible patterns of symptom presentation – either unifocal bone pain affecting the clavicle or multifocal pain involving the lower limb.

### Investigations

In our cohort, a lower percentage of patients underwent bone biopsies (51.1%) compared to the German National Surveillance Study [7] (61%), whereas a higher proportion had whole-body MRI (80% versus 60%). MRI imaging is thought to be more sensitive than other imaging modalities at identifying asymptomatic lesions [14]. While 57% of our cohort had reported multifocal bone pain at presentation, a study by Andronikou *et al.* [15] using whole-body MRI analysis in a cohort of 37 patients demonstrated multifocal disease in 89% of patients. Chronic inflammation and fibrosis were the most common descriptions in the bone biopsy reports, a finding consistent with that of Bhat *et al.* [5].

### Initial Management

To date, there has not been any randomised controlled trial to determine the optimal treatment of CRMO. In line with previous studies, NSAIDs are the predominant first-line treatment in this cohort, given to 93.9% of patients. The second most common treatment is bisphosphonates (44.8%). Although there are relatively few patients treated within the first few months by steroids, biologics or methotrexate, this number may increase during follow up. We found that younger patients and those with higher CRP were more likely to receive antibiotics. This may be attributed to potential diagnostic uncertainties in younger patients and exercising clinical caution in patients with higher CRP levels.

### Healthcare Utilisation

This is the first study assessing healthcare utilisation in CRMO. While most patients were diagnosed by rheumatologists, 17.4% were diagnosed by orthopaedic specialists, 9.8% by general paediatricians and 1.6% by maxillo-facial surgeons. This underscores the importance of raising awareness among health professionals in these specialties. The significant number of allied health professional referrals, inpatient admissions and emergency department visits highlight the necessity for appropriate funding and resource allocation to ensure optimal care for children and young people with CRMO. Furthermore, our data on the functional impact, including school attendance (only 87.2% in full attendance) and mobility (33% with impaired mobility), underscores the significant impact of this condition on children and their families.

### Conclusion and Future Direction

CRMO is a rare auto-inflammatory disorder that continues to be a challenge to investigate, diagnose, and manage. The disease can present with a range of clinical features in its early phase. This study represents the most comprehensive surveillance effort to date on CRMO, revealing an estimated incidence of 0.65 per 100,000 children per year. A notable sex difference is observed, with a 2:1 female:male preponderance. Male patients are more likely to present with multifocal disease, and there appears to be two distinct disease patterns with either clavicular or leg pain presentations. Our cohort had a shorter diagnostic delay (5 months) and a high proportion of whole-body MRI utilisation compared to previous studies.

Our future plans include analysis of the MRIs in the cohort by an expert musculoskeletal radiology panel to identify key radiological features of CRMO. The ongoing two-year follow-up of the cohort will provide insights into the medium-term treatment and outcomes. Together with this study, these results will serve as a crucial baseline for informing clinical care guidelines and guiding future research focused on children and young people grappling with CRMO.

## Data Availability

All data produced in the present study are available upon reasonable request to the corresponding authors.

## Data Availability Statement

The data underlying this article will be shared on reasonable request to the corresponding author.

## Acknowledgements

We gratefully acknowledge Clare Lyon for her invaluable administrative support in facilitating this study. Additionally, we are profoundly grateful for the support and valuable insights contributed by our patient advisory group, consisting of three parent representatives of affected children, the CRMO-UK Facebook group, and the Patient and Public Involvement team from NIHR Cambridge BRC.

## Supplement

### Supplementary S1A: Analytic Case Definition

Bristol diagnostic criteria [2] (wording has been modified for clarification):

- The presence of localised bone pain, which could be single site or multiple sites

AND

- The presence of the following radiological findings on plain X-ray: lytic areas, sclerosis or new bone formation, or on MRI: bone marrow oedema, or periostitis.

AND EITHER

- Criterion 1: More than one bone (or clavicle alone) with CRP < 30 g/L.OR

OR

- Criterion 2: Bone biopsy obtained whilst not on antibiotic therapy shows inflammatory changes (plasma cells, osteoclasts, fibrosis or sclerosis) and no bacterial growth.

**Supplementary S1B:**
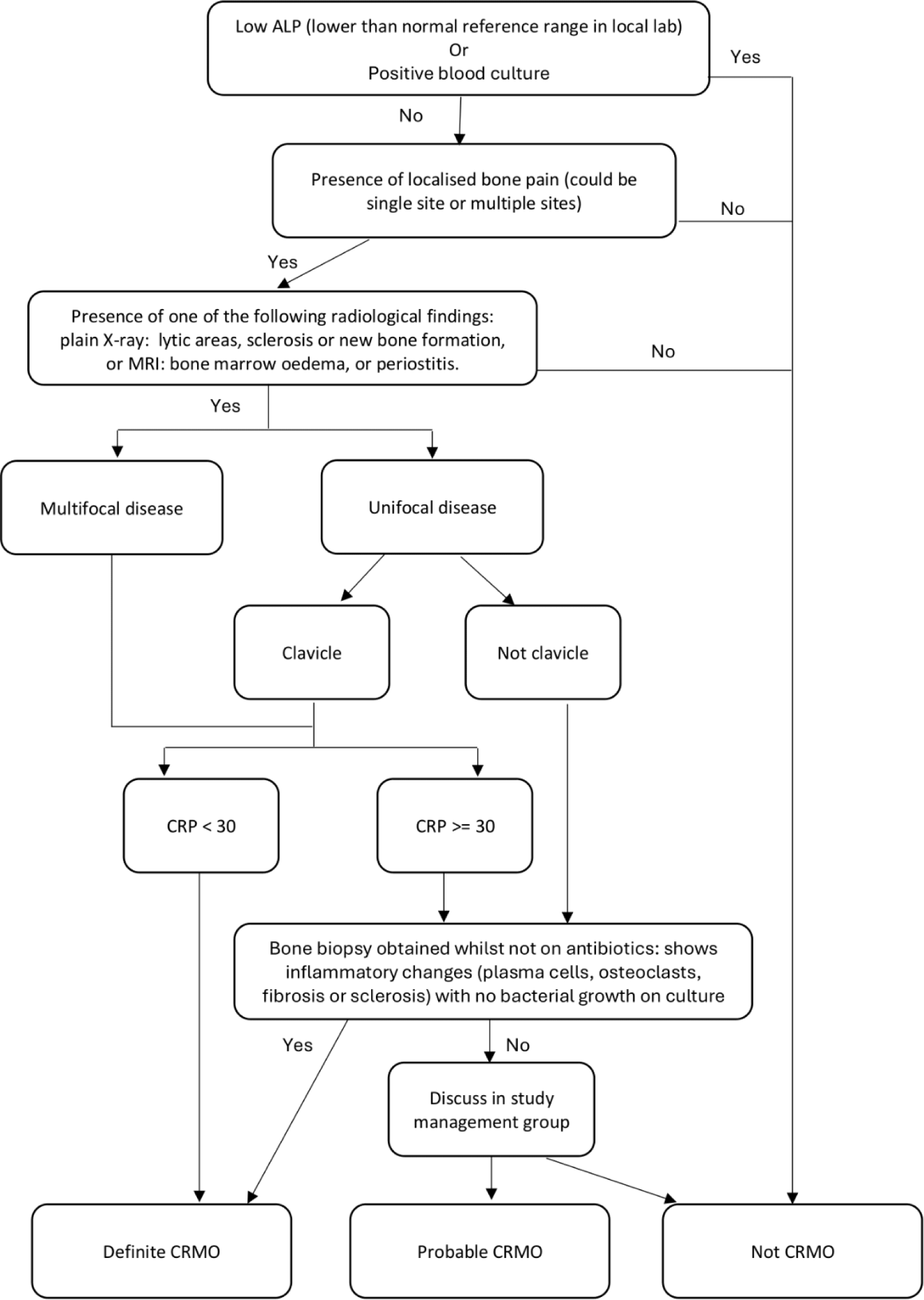
Decision Tree for Case Inclusion / Exclusion.

**Supplementary Figure S2:**
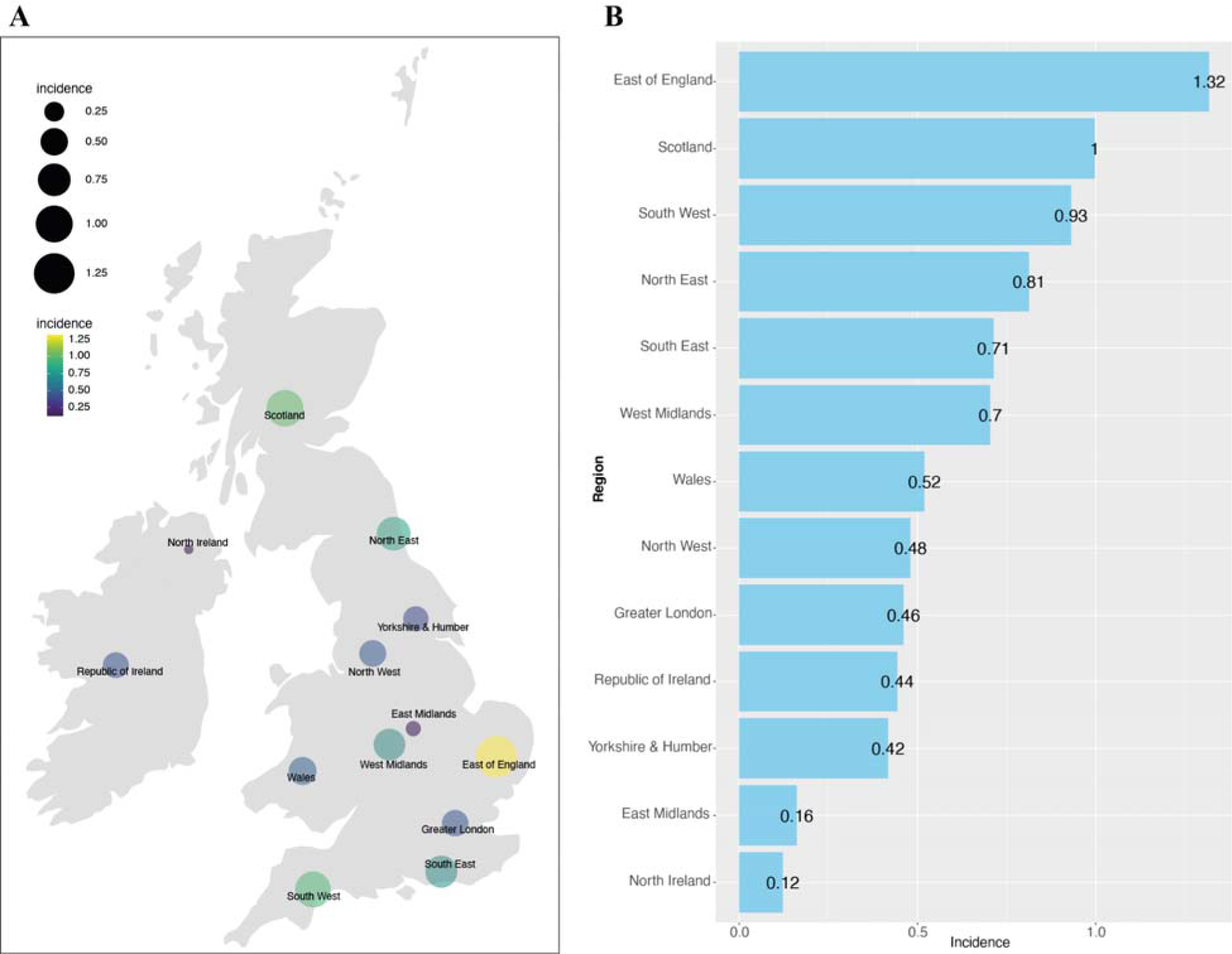
CRMO incidences by nation & region. (A) Incidences of CRMO (per 100,000 children) by nation and region on the map of UK and ROI. (B) Bar chart showing the incidences of CRMO (per 100,000 children) by nation and region in decreasing order.

**Supplementary Figure S3:**
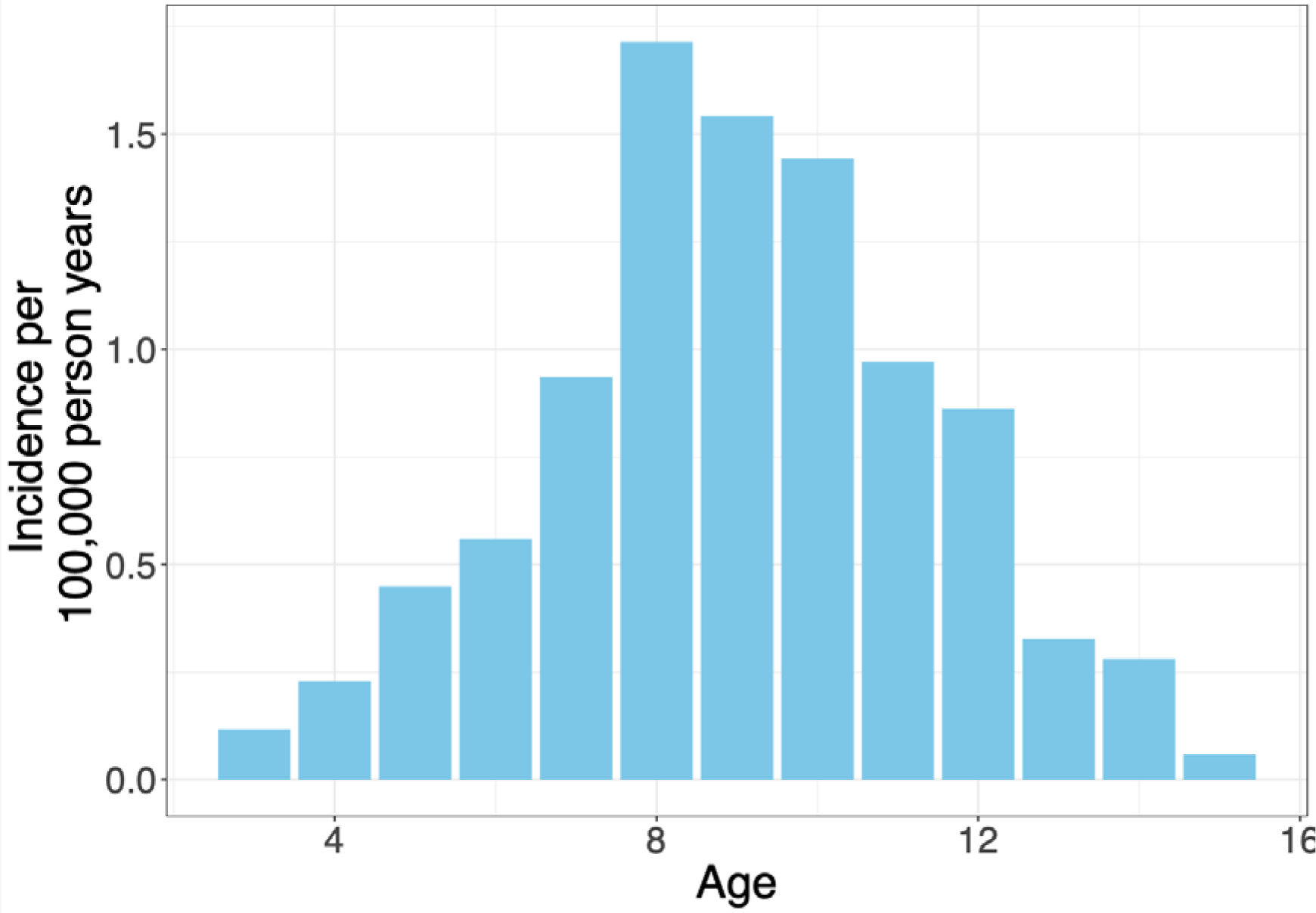
Bar chart of CRMO incidences by age.

**Supplementary Figure S4:**
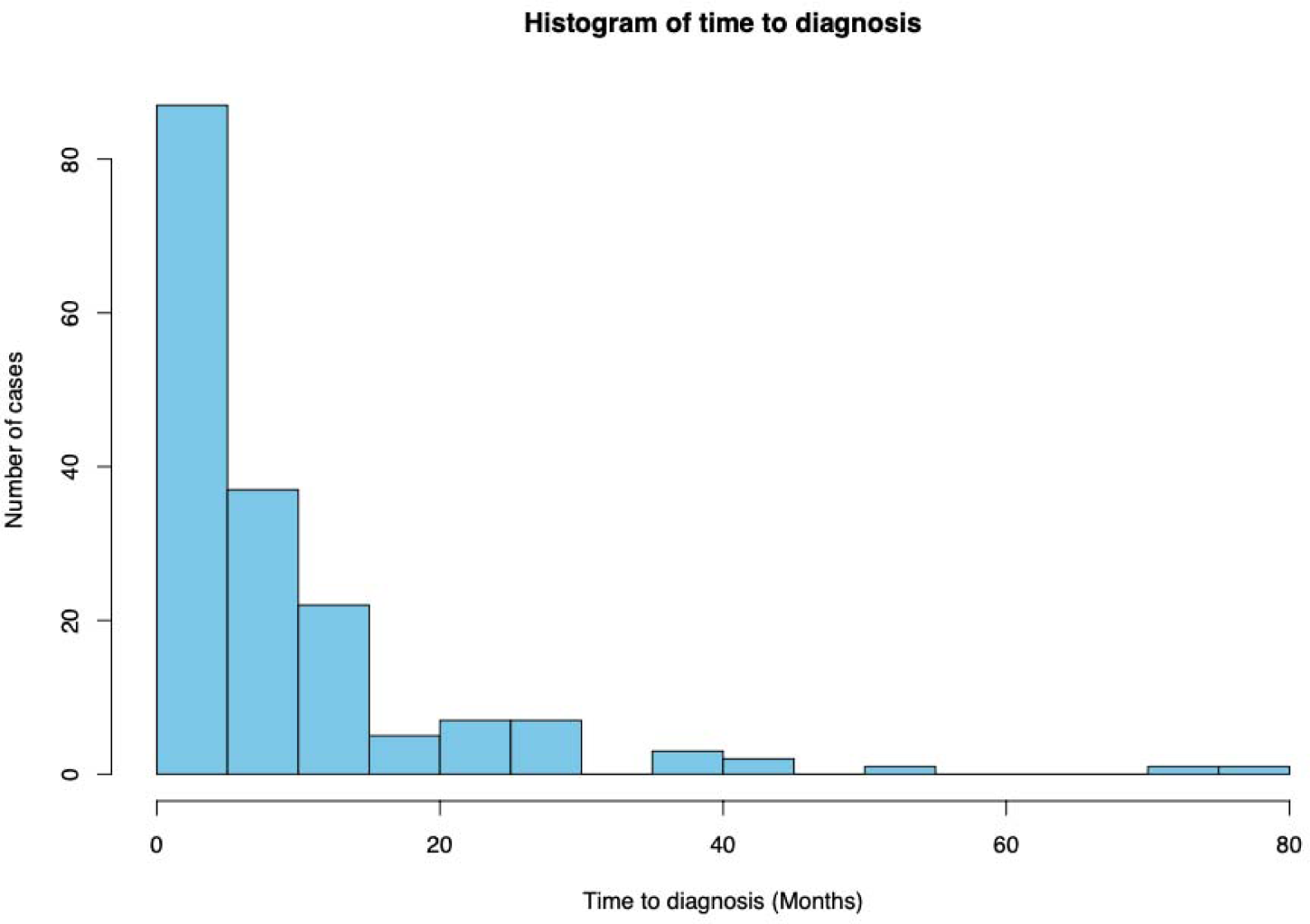
Histogram of time to diagnosis.

**Supplementary Figure S5:**
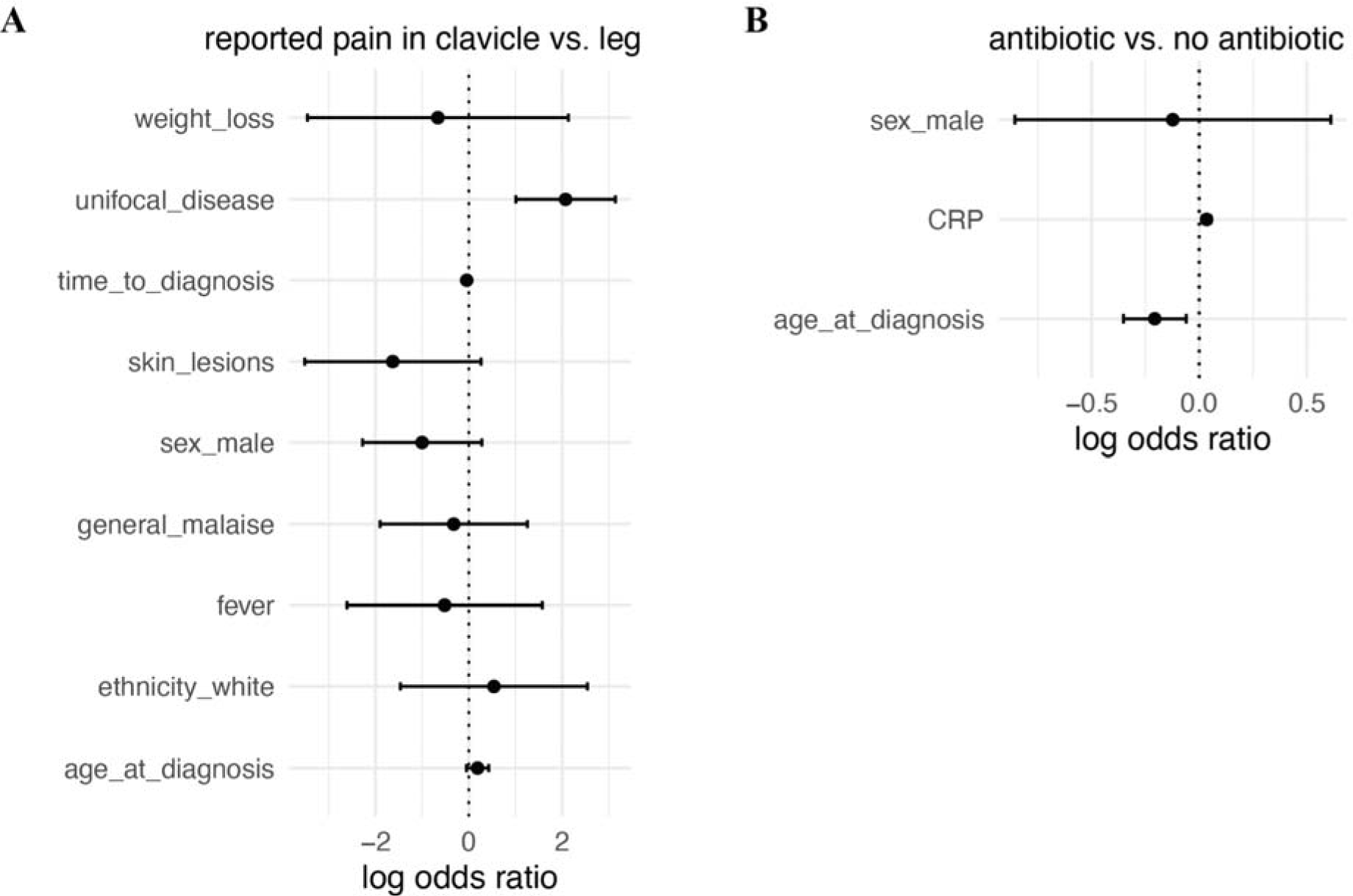
Multivariate logistic regression analyses. (A) Results of multivariate logistic regression analysis on reported pain in clavicle versus leg. The dots represent the estimated mean of log odds ratio, with the error bars representing 95% confidence intervals. Cases with missing data in any of the tested covariates were excluded, and only cases with reported bone pain in either clavicle or leg are included (n = 101). (B) Results of multivariate logistic regression analysis on associations with antibiotic treatment. The dots represent the estimated means of log odds ratio, with the error bars representing 95% confidence intervals. Cases with missing data in any of the tested covariates were excluded, and 170 cases were included in the analysis.

**Supplementary Figure S6:**
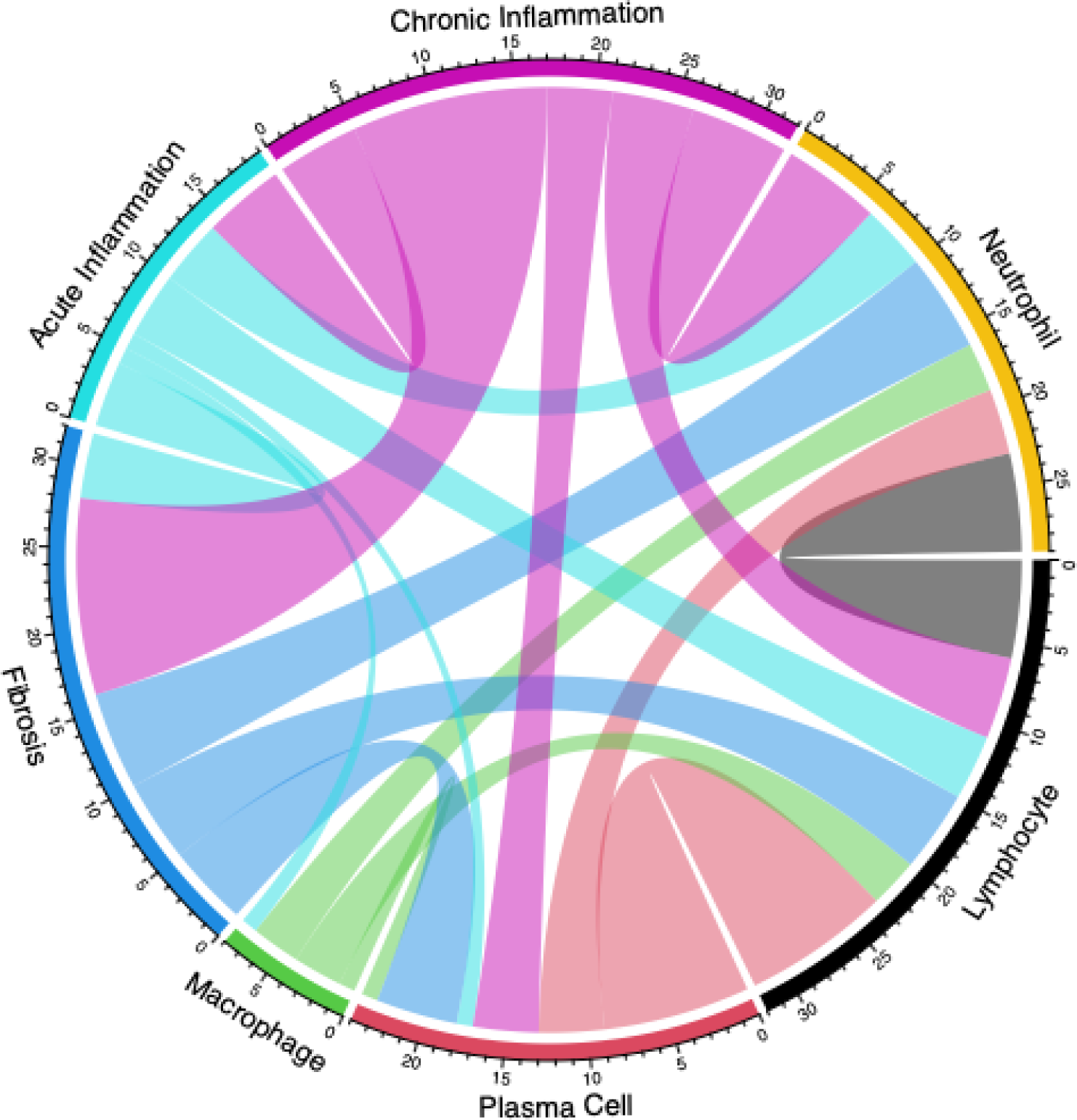
Chord diagram showing pairwise co-occurrences of the histopathology feature.

## References

1 Giedion A, Holthusen W, Masel LF, et al. [Subacute and chronic ‘symmetrical’ osteomyelitis]. Ann Radiol. 1972;15:329–42.

2 Roderick MR, Shah R, Rogers V, et al. Chronic recurrent multifocal osteomyelitis (CRMO) - advancing the diagnosis. Pediatr Rheumatol Online J. 2016;14:47.

3 Girschick H, Finetti M, Orlando F, et al. The multifaceted presentation of chronic recurrent multifocal osteomyelitis: a series of 486 cases from the Eurofever international registry. Rheumatology. 2018;57:1203–11.

4 Walsh P, Manners PJ, Vercoe J, et al. Chronic recurrent multifocal osteomyelitis in children: nine years’ experience at a statewide tertiary paediatric rheumatology referral centre. Rheumatology. 2015;54:1688–91.

5 Bhat CS, Anderson C, Harbinson A, et al. Chronic non bacterial osteitis-a multicentre study. Pediatr Rheumatol Online J. 2018;16:74.

6 Wipff J, Costantino F, Lemelle I, et al. A large national cohort of French patients with chronic recurrent multifocal osteitis. Arthritis Rheumatol. 2015;67:1128–37.

7 Jansson AF, Grote V, ESPED Study Group. Nonbacterial osteitis in children: data of a German Incidence Surveillance Study. Acta Paediatr. 2011;100:1150–7.

8 BPSU impact - case studies. RCPCH. https://www.rcpch.ac.uk/resources/bpsu-impact-case-studies (accessed 5 December 2023)

9 Ethnicity of children aged 5 to 16 in UK regions, 2020. 2021. https://www.ons.gov.uk/peoplepopulationandcommunity/culturalidentity/ethnicity/adhocs/13217ethnicityofchildrenaged5to16inukregions2020 (accessed 5 December 2023)

10 Springate DA, Parisi R, Kontopantelis E, et al. Incidence, prevalence and mortality of patients with psoriasis: a U.K. population-based cohort study. Br J Dermatol. 2017;176:650–8.

11 Aden S, Wong S, Yang C, et al. Increasing Cases of Chronic Nonbacterial Osteomyelitis in Children: A Series of 215 Cases From a Single Tertiary Referral Center. J Rheumatol. 2022;49:929– 34.

12 Borzutzky A, Stern S, Reiff A, et al. Pediatric chronic nonbacterial osteomyelitis. Pediatrics. 2012;130:e1190–7.

13 Kaiser D, Bolt I, Hofer M, et al. Chronic nonbacterial osteomyelitis in children: a retrospective multicenter study. Pediatr Rheumatol Online J. 2015;13:25.

14 Khanna G, Sato TSP, Ferguson P. Imaging of chronic recurrent multifocal osteomyelitis. Radiographics. 2009;29:1159–77.

15 Andronikou S, Mendes da Costa T, Hussien M, et al. Radiological diagnosis of chronic recurrent multifocal osteomyelitis using whole-body MRI-based lesion distribution patterns. Clin Radiol. 2019;74:737.e3–737.e15.

